# PSMA-Hornet: fully-automated, multi-target segmentation of healthy organs in PSMA PET/CT images

**DOI:** 10.1101/2022.02.02.22270344

**Authors:** Ivan S. Klyuzhin, Guillaume Chaussé, Ingrid Bloise, Juan Lavista Ferres, Carlos Uribe, Arman Rahmim

## Abstract

Prostate-specific membrane antigen (PSMA) PET imaging represents a valuable source of information reflecting disease stage, response rate, and treatment optimization options, particularly with PSMA radioligand therapy. Quantification of radiopharmaceutical uptake in healthy organs from PSMA images has the potential to minimize toxicity by extrapolation of the radiation dose delivery towards personalization of therapy. Furthermore, factors affecting biodistribution of PSMA radiotracers that remain mostly unknown can be investigated by analyzing PSMA PET images with segmented organs. However, segmentation and quantification of uptake in organs requires labor-intensive organ delineations that are often not feasible in the clinic nor scalable for large clinical trials. In this work we have developed and tested the PSMA Healthy organ segmentation network (PSMA-Hornet), a fully-automated deep neural net for effective and robust segmentation and labelling of 14 healthy organs representing the normal biodistribution of [^18^F]DCFPyL on PET/CT images.

**Methods:** The study used manually-segmented [^18^F]DCFPyL PET/CT images from 100 subjects, and 526 similar images without segmentations. The unsegmented images were used for self-supervised model pretraining. For supervised training, 3-fold cross-validation was used to evaluate the network performance, with 85 subjects reserved for model training, 5 for validation, and 10 for testing. Image segmentation and quantification metrics were evaluated on the test set with respect to manual segmentations by a nuclear medicine physician.

**Results:** With our best model, the lowest mean Dice coefficient on the test set was 0.826 for the sublingual gland, and the highest was 0.964 for liver. The highest mean error in tracer uptake quantification was 13.9% in the sublingual gland. Self-supervised pretraining improved training convergence, train-to-test generalization, and segmentation quality. In addition, we found that a multi-target network produced significantly higher segmentation accuracy than single-organ networks.

**Conclusion:** The developed network can be used to automatically obtain high-quality organ segmentations for PSMA image analysis tasks. It can be used to reproducibly extract imaging data, and holds promise for clinical applications such as personalized radiation dosimetry and improved radioligand therapy.

## INTRODUCTION

Prostate cancer is the most common solid tumor and the second most frequent cancer diagnosed in men (*1*). After radiation therapy and/or prostatectomy is applied to the primary tumor, up to 30% of patients develop recurrence with possible metastases (*2*). Metastatic, castration-resistant prostate cancer (mCRPC) leads to the death of over 350K people globally every year (*3, 4*). Overexpression of prostate-specific membrane antigen (PSMA) in prostate cancer cells is correlated with more advanced and progressive disease (*5*). Thus, PSMA-targeting radioligand therapy (PRLT), and its imaging counterpart, represent a new promising frontier in prostate cancer treatment (*6, 7*).

At present, the de-facto standard of care is to administer a fixed dose of PRLT to every patient. However, it has been shown that for healthy tissues, the absorbed doses between patients can vary on the order of 10–20 folds (*8, 9*). The fixed-dose approach uses conservative administered activities to minimize the risk of organ damage across the entire population, which results in a large number of under-treated patients. Early evidence shows that PSMA PET imaging can be used to estimate the impact of PRLT on healthy organs prior to the beginning of treatment, thus allowing a more tailored and safely maximized drug delivery to each patient. For example, in a study by Violet et al. (*9*), significant correlation was found between pre-therapy PSMA PET standardized uptake values (SUVs) in healthy organs and absorbed dose during therapy. PSMA PET images can likewise be used to estimate the total tracer uptake in mCRPC lesions, which has been shown to correlate with outcomes (*10, 11, 12*). However, the use of PSMA PET/CT images for these applications is hindered by the need to manually segment organs at risk, which is an operator-dependent and very labor-intensive process.

In addition to potential applications in dosimetry, PSMA PET images represent a valuable source of information that can be linked to cancer prognosis, response rate, and choice of therapy regimen (*13*). Automated segmentation of organs would allow the mining of large imaging datasets for such hidden data.

In this work, we have developed the PSMA Healthy organ segmentation network (PSMA-Hornet), an artificial neural net for fully-automated segmentation of 14 organs with high tracer uptake on [^18^F]DCFPyL PET/CT images. To develop and train the model, we utilized a novel self-supervised pretraining method, which allowed us to leverage a large number of unsegmented PET/CT images to improve the training convergence and model performance. The network has customized convolutional architecture that includes batch normalization layers and a multi-slice/multi-modality fusion block. With the use of careful training batch balancing, we trained the model to segment 14 organs simultaneously, which improved the segmentation quality compared to single-organ models.

We compare our model-produced segmentations with organ delineations by a nuclear medicine physician. We first report Dice coefficients measured for a baseline model, and for models with incrementally added custom modifications (batch normalization, pretraining, and multi-target training). Second, we report recall, precision, and Hausdorff distance for single-organ and multi-organ networks. Finally, we compare several image metrics in different organs quantified according to manual and automated segmentations.

## METHODS

### Data description

The study used [^18^F]DCFPyL PET/CT images from 100 subjects participating in a local PSMA PET clinical trial (NCT02899312) that were clinically reported as lesion-negative. In addition, images of 526 lesion-positive and negative subjects from the same trial were used for self-supervised network pretraining, as elaborated below. The collection of imaging data was approved by the University of British Columbia - BC Cancer Ethics Board (UBC BC Cancer REB), and all subjects gave informed written consent. Study participants were injected intravenously with approximately 350 MBq of [^18^F]DCFPyL (with adjustment for body weight) after a 4-hour fast. After a 120-minute uptake period, the subjects were scanned on a GE Discovery 600 or 690 scanner (GE Healthcare, USA). The PET transaxial image size was 192×192 voxels (voxel size 3.64 mm), and the CT image size was 512×512 voxels (voxel size 1.36 mm). The axial PET/CT image extent was either 268 or 300 planes, depending on the subject’s height. The representative subject demographics and inclusion criteria were published in a previous study (*14*).

### Image segmentation

On the set of 100 PET/CT images, organs with physiologic tracer uptake were segmented by a nuclear medicine physician (GC): lacrimal glands (x2), parotid glands (x2), submandibular glands (x2), tubarial gland, sublingual gland, spleen, liver, kidneys (x2), bowel, and bladder (Fig. 1). Contours around organs were drawn using commercially available software (MIM Software, USA). First, using non-zero voxels, the contour of the patient was drawn. A fixed SUV threshold excluded voxels below SUV 5. Non-connected regions were then split into individual regions of interest and manually verified. Subsegmentation of overlapping contours was done manually by drawing sector lines to split contours into sections and by running a cluster analysis tool. Regions that fell in part or entirely below the SUV 5 threshold but still had above-background uptake were added by manually with the help of an edge-detection gradient algorithm (PETedge). Labels were attributed by an in-house developed algorithm based on the relative locations of the contours. The outcome of labeling process was visually verified and manually corrected where needed. In those cases where no above-background uptake was present on PET images, the corresponding organ was not segmented; an empty circular 2D mask was used as a placeholder to allow the labelling algorithm to detect and label all regions appropriately. This semi-automated workflow with manual interventions and corrections was validated in another study (*15*).

**FIGURE 1:**
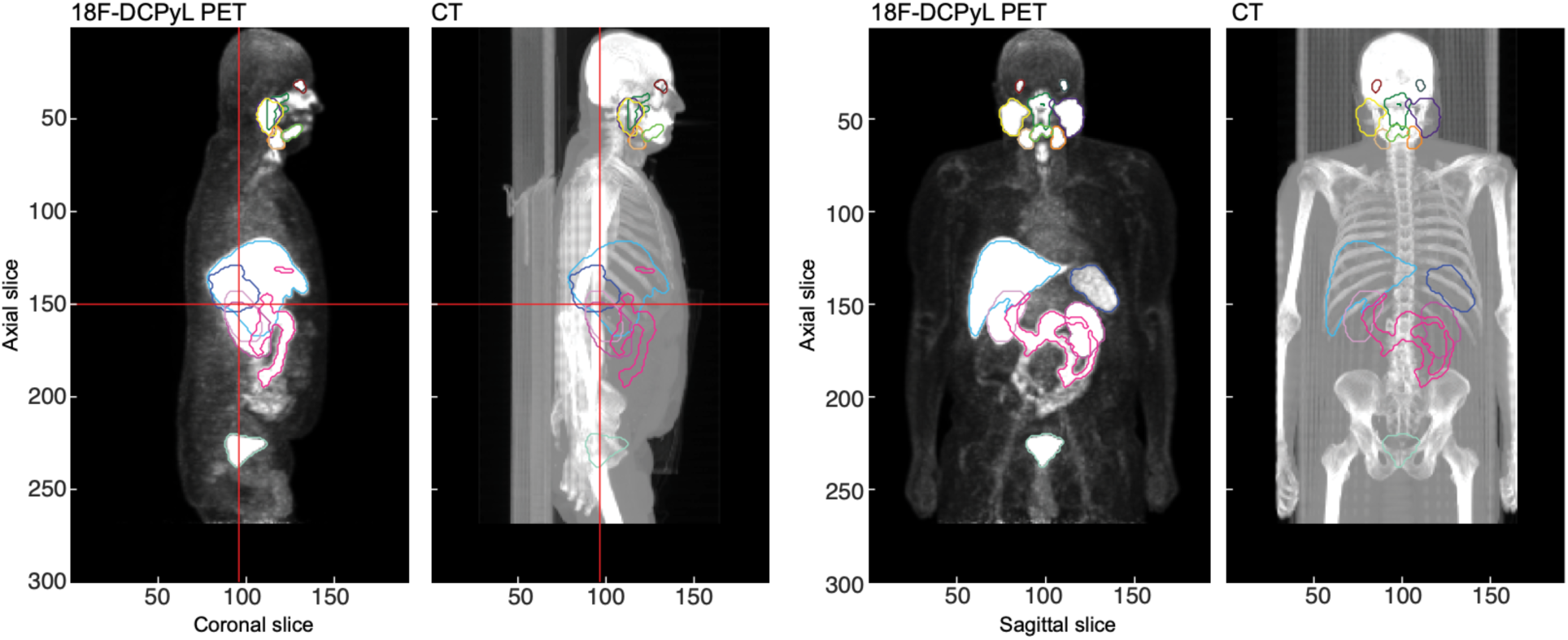
Examples of physician-produced delineations of 14 organs for a subject with negative mCRPC findings. (*left*) PET and corresponding CT images (maximum intensity projections) in sagittal view, and (*right*) same images in the coronal view.

### Pre-processing of images and labels

After the acquisition, CT images were down-sampled to PET voxel size using slice-by-slice 2D bilinear interpolation and cropped to PET image size in a way to preserve PET/CT alignment. Both PET and CT images were normalized to take on voxel values approximately within 0–1 range: the PET SUV values were divided by 10, while CT Hounsfield units (HU) in the range from 800 to 1200 (i.e., a window level of 1000 HU with a window width of 400 HU) were linearly mapped to [0, 1] in order to enhance the contrast for soft tissues. Healthy organ segmentations originally stored as contours in RTSTRUCT files were converted to binary 3D masks with the exact image dimensions as PET and CT images. Bilateral organs were combined into a single class (e.g., left kidney and right kidney were labeled together as “kidneys”), producing a total of 10 targets and background for automated segmentation.

### PSMA-Hornet architecture

The fully-convolutional architecture of PSMA-Hornet is based on the U-net with several modifications (Fig. 2A). The network is based on the U-net fully-convolutional architecture with the following modifications. The network has two inputs represented by axial PET and CT slices; to provide contextual information, neighboring slices are also supplied in separate channels (i.e., 3 PET and 3 CT slices). Further, a custom modality-fusion block was introduced at the input, inspired by the inception modules (*16*). The block passes the images from two modalities through different convolutional pathways with kernel sizes 1×1, 3×3, 5×5, and 7×7. The multi-kernel feature maps are concatenated, and another convolution is taken over the combined 64 feature maps. The rationale for this design, is that PET and CT images have different noise levels and scales of anatomical/functional landmarks. Separate convolutional pathways allow the low-level CT features to not be affected by (higher) PET noise, while convolutional kernels of different sizes allow for learning of features on different scales (*17*).

**FIGURE 2:**
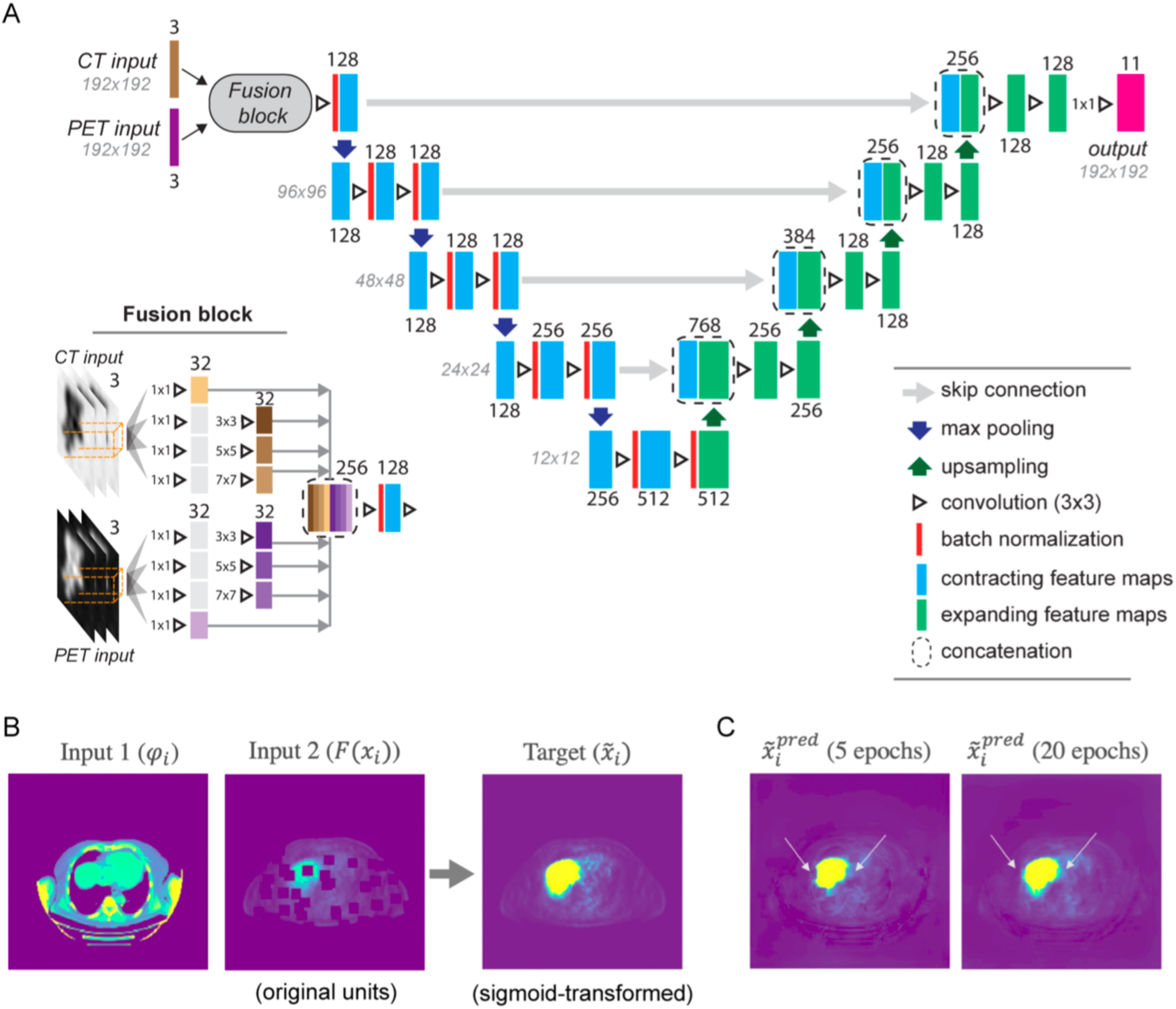
A. Diagram of the PSMA-Hornet architecture. Compared to a standard U-net, batch normalization layers were added throughout the contracting path, dropout layers were removed, a custom multi-slice modality fusion block was added to the input, and the network had multiple targets (outputs). B. The self-supervised pretext task was to predict a sigmoid-transformed PET slice, from a pair of CT and PET slices. Information was partially removed from the input PET image by deleting random image patches. C. In the process of self-supervised pretraining, the network learned to correctly predict organ boundaries.

To improve the gradient propagation and training convergence, batch normalization layers were added after each convolutional layer on the contracting path of the network (*18*). A version of the network was tested where batch normalization was added to both contracting and expanding paths, however this worsened the training convergence compared to modifying only the contracting path. Dropout layers were not used as they were found to be detrimental for training convergence and segmentation quality on the validation set. The test set(s) were not used when exploring these architecture modifications.

The output of PSMA-Hornet was represented by a layer with 11 probability maps, 10 for the target organs and one for the background. The multi-label segmentation image was obtained by choosing the class with the highest probability for each voxel. All layers in the network had a leaky ReLU activation function to alleviate the vanishing gradient problem, and the output layer had sigmoid activation to restrict the output values to the range [0, 1] (for probability-based interpretation). The network was implemented in Python v.3.6 using TensorFlow v.2.2, and had 9,373,003 trainable parameters in total, or 937,300 per target.

### Self-supervised pretraining

In addition to 100 manually-segmented PSMA PET/CT images, we had access to 526 images without organ segmentations. PSMA-Hornet was pretrained on this larger dataset in a self-supervised manner, using the context-recovery pretext task (*19*). Given a CT image slice *𝝋* and (unlabeled) PET image slice ***x***, in this task *G* we trained the network to predict the sigmoid-transformed PET image slice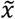:

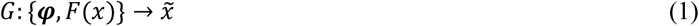

Here *F*(⋅) is an augmentation function that removed (i.e., set to zero) random patches in the input PET slice (Fig. 2B). The patches were 11×11 voxels in size, and a total of 50 patches were removed per slice. The target PET image was sigmoid-transformed using the equation:

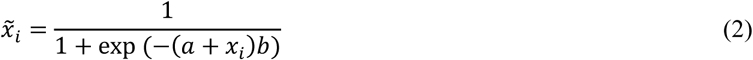

where *x*_*i*_ are the original voxel values, and 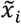 are the transformed values. We used *a* = -2 and *b* = 2 so that background SUV values mapped approximately to 0, and organ SUV values mapped approximately to 1. The rationale is that images transformed in such a manner have approximately the same appearance as binary masks, thus making the self-supervised pretext task very similar to supervised segmentation learning. We used the mean squared error loss for pretraining:

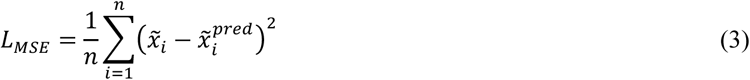

where 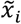 are the true voxel values, 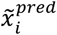 are the predicted values, and *n* is the total number of voxels in a batch. In the pre-training phase, the output layer of the network only contained one feature map.

In the process of pretraining, the network progressively learned to fill in the missing image matches from contextual information. This process is illustrated in Fig. 2C, where patches were removed from several locations in the liver and surrounding tissues. With increasing training epochs, the network learned to better predict the anatomical shape of the liver (as indicated by arrows).

### Supervised training

Following self-supervised pretraining, PSMA-Hornet was trained in a supervised manner using the 100 PET/CT patient scans with manual organ segmentations. The data were split into 3 folds, and in each fold 85 scans were randomly selected for training, 5 for validation, and 10 for testing. The total number of 2D slices available for training in each fold was approximately 32,300. We found that it was necessary to account for class imbalance for the training to reliably converge. Since PSMA-Hornet uses 2D image slices as inputs, there are two types of class imbalance that need to be addressed.

The first type is the in-plane class imbalance originating from different image areas occupied by different organs. In addition, all organs are small with respect to the background, which was treated as a separate class. To mitigate this we used the soft Dice loss function (*20*) defined for a single target by the equation:

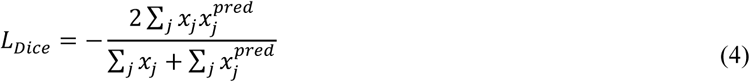

where *x*_*j*_ is the true voxel value (binary), and 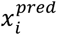 is the predicted voxel value (sigmoid). The sum is taken across all voxels in the training batch. To extend this loss to multi-target training of PSMA-Hornet, it was re-defined as:

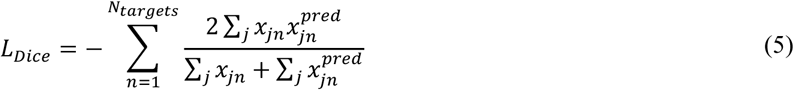

where *x*_*jn*_ is the true voxel value for target/organ *n*, and 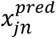 is the predicted voxel value. The lowest possible value of the multi-class Dice loss is -*N*, where *N* is the number of targets.

The second type of class imbalance is due to the varying axial extent of organs, which resulted in drastically different number of training samples available for different organs. For example, the sublingual gland was present in 620 planes (taken from different subjects), while the bowel was present in 4094 planes. To address this imbalance, each training batch of 64 samples was constructed to have an equal representation of all organs (in terms of the number of samples containing those organs). Thus, organs with small axial extent were proportionally oversampled.

The training loss was minimized using the Adam algorithm with learning rate set to 10^−5^. Training was performed on a Microsoft Azure virtual machine (NC24s v3 series) with 4 Tesla V100 GPUs (16 Gb), and parallelized by splitting each training batch between the GPUs, i.e. 16 training samples per GPU. Training was performed for 200 epochs, which were verified to be sufficient for minimization of training and validation losses.

### Performance analysis

The network was trained, validated, and tested on 3 folds. The test samples from each fold were combined, totaling 30 samples, and model performance metrics were computed on this combined set. We measure several metrics of segmentation quality as well as “downstream” image quantification metrics.

To assess the segmentation quality, we computed the Dice similarity coefficient, recall, precision and Hausdorff distance for each target organ. All metrics were computed with respect to organ delineations produced by a nuclear medicine physician. To provide a reference point, we compare the Dice coefficients to inter-rater agreement that was measured in a previous study with three human operators (*15*).

Quantitative “downstream” image metrics included:

- mean absolute error (MAE) of total organ volume,
- MAE of mean SUV (SUV_mean_) in the organ,
- MAE of total uptake (TU), computed as an integral of SUVs over the segmentation volume.

All metrics were computed using the equation

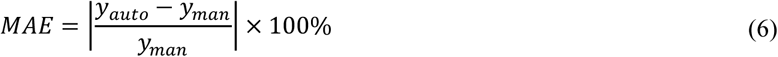

where *y*_*man*_ is a metric computed using manual segmentations (volume, SUV_mean_, or TU) and *y*_*auto*_ is that metric computed using automated segmentations from PSMA-Hornet.

## RESULTS

### Baseline networks and effect of pretraining

We first compare segmentation quality between single-organ networks: standard U-net, and networks with added batch normalization and self-supervised pre-training. The Dice coefficients for these networks, measuring the agreement between manual and automatic segmentations on the test sets, are given in Table 1 (the best values in each row are highlighted in bold); all networks had single-slice inputs. The data show that the addition of batch normalization improved Dice coefficients for all organs. The most substantial improvements were found for the sublingual (0.408 to 0.745) and tubarial (0.627 to 0.798) glands. Both of these organs were likely difficult to segment for the baseline model due to their small size and low SUV values (SUV_mean_ for both organs was approximately 3.0). The consistency of segmentations was also improved for all organs, as demonstrated by lower standard deviations of the Dice coefficients.

**TABLE 1:**
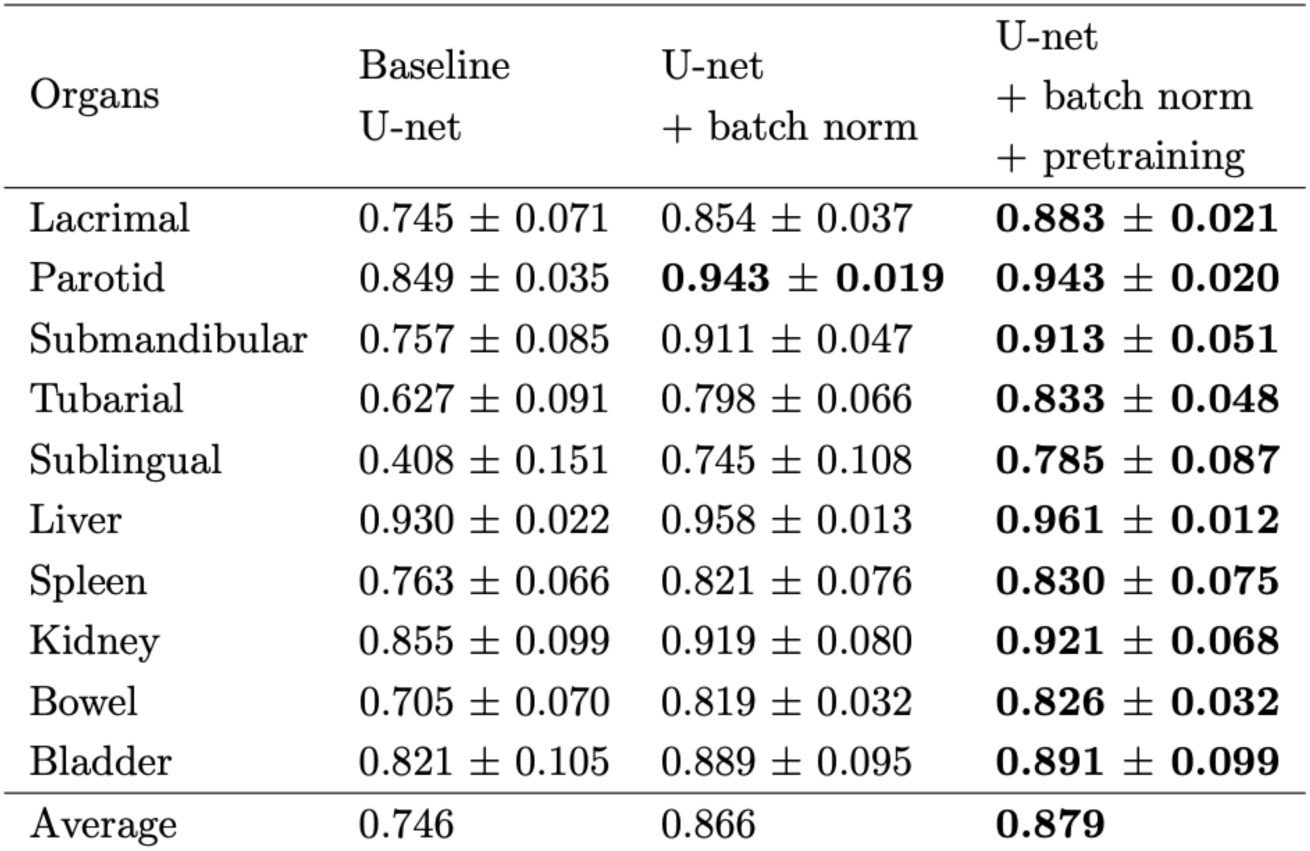
Dice coefficients measured with single-slice, single-organ networks.

| Organs | Baseline<br>U-net | U-net<br>+ batch norm | U-net<br>+ batch norm<br>+ pretraining |
| --- | --- | --- | --- |
| Lacrimal | $0.745 \pm 0.071$ | $0.854 \pm 0.037$ | **$0.883 \pm 0.021$** |
| Parotid | $0.849 \pm 0.035$ | **$0.943 \pm 0.019$** | **$0.943 \pm 0.020$** |
| Submandibular | $0.757 \pm 0.085$ | $0.911 \pm 0.047$ | **$0.913 \pm 0.051$** |
| Tubarial | $0.627 \pm 0.091$ | $0.798 \pm 0.066$ | **$0.833 \pm 0.048$** |
| Sublingual | $0.408 \pm 0.151$ | $0.745 \pm 0.108$ | **$0.785 \pm 0.087$** |
| Liver | $0.930 \pm 0.022$ | $0.958 \pm 0.013$ | **$0.961 \pm 0.012$** |
| Spleen | $0.763 \pm 0.066$ | $0.821 \pm 0.076$ | **$0.830 \pm 0.075$** |
| Kidney | $0.855 \pm 0.099$ | $0.919 \pm 0.080$ | **$0.921 \pm 0.068$** |
| Bowel | $0.705 \pm 0.070$ | $0.819 \pm 0.032$ | **$0.826 \pm 0.032$** |
| Bladder | $0.821 \pm 0.105$ | $0.889 \pm 0.095$ | **$0.891 \pm 0.099$** |
| Average | 0.746 | 0.866 | **0.879** |

The addition of self-supervised pre-training further improved the Dice coefficients and their standard deviations for all organs except for parotid glands. Importantly, pre-training turned out to be critically important for larger networks with multi-slice inputs and multi-target outputs. The effect of pre-training on the network training convergence is illustrated in Fig. 3, where (supervised) training and validation losses are plotted as functions of training epoch. The plots demonstrate that without pre-training, there was a relatively large difference between training and validation losses (i.e. large generalization error), and the validation loss tended to be unstable. Self-supervised pre-training improved the rate of loss minimization and also made the validation loss more stable and monotonic.

**FIGURE 3:**
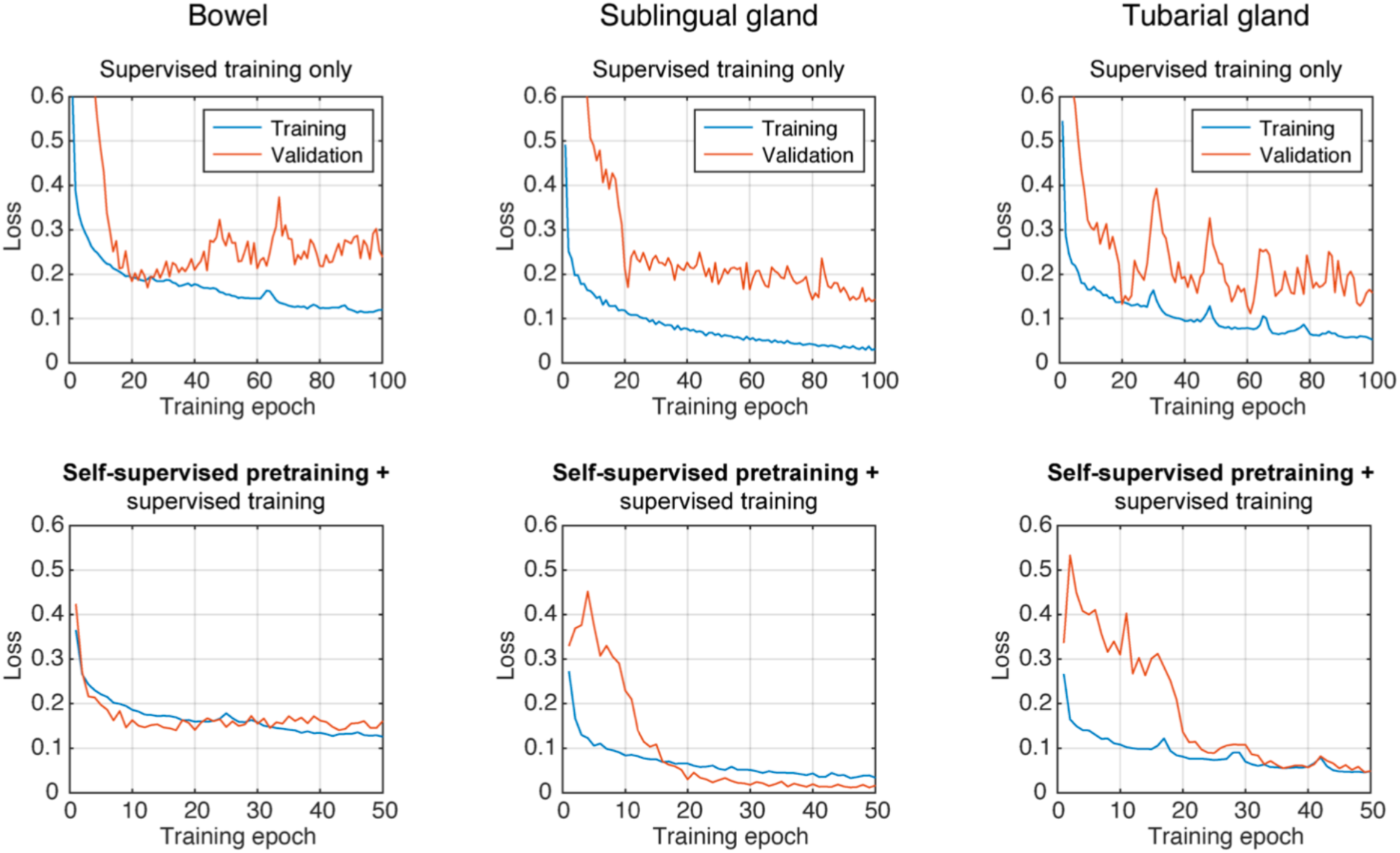
Plots of training and validation losses versus the training epoch for three organs: bowel (left), sublingual gland (middle), and tubarial gland (right). Top row, training of a 3-slice single-target U-net from scratch. Bottom row, training of a similar network that had been pre-trained on unlabeled data. The networks included batch normalization. Note the difference of scales on the x-axis.

The substantial increase in network performance with batch normalization and self-supervised pre-training was observed for multiple organs, increasing the confidence that the improvements were real and did not occur by random chance. We also determined that, even with simpler baseline networks, the use of in-batch class balancing via minority class oversampling was necessary for reliable training convergence, particularly with small organs like lacrimal glands.

### Multi-slice, multi-target networks

Compared to the networks that used a single PET and CT slice as inputs, using the multi-slice input with modality fusion block improved the Dice coefficients for lacrimal glands, tubarial gland, sublingual gland, and bladder (Table 2, first column). Notably, it significantly reduced spurious false positives that only had one-slice thickness in the axial dimension. We had chosen to keep the multi-slice input and the modality fusion block in the final model since it improved the segmentation for the sublingual gland, an organ at high risk that otherwise had the lowest Dice score (0.785).

**TABLE 2:** Dice coefficients obtained with single-organ and multi-organ networks, with reference to inter-rater agreement. The single-target networks had 1,960,577 trainable parameters in total, thus twice as many per-target compared to PSMA-Hornet.

| Organs | Single-organ<br>networks | Multi-organ<br>network<br>(PSMA-Hornet) | Human inter-rater<br>agreement |
| --- | --- | --- | --- |
| Lacrimal | $0.898 \pm 0.027$ | **$0.912 \pm 0.016$** | $0.893 \pm 0.017$ |
| Parotid | $0.948 \pm 0.021$ | **$0.950 \pm 0.016$** | $0.936 \pm 0.014$ |
| Submandibular | $0.909 \pm 0.038$ | **$0.933 \pm 0.026$** | $0.907 \pm 0.015$ |
| Tubarial | $0.841 \pm 0.072$ | **$0.874 \pm 0.038$** | $0.788 \pm 0.028$ |
| Sublingual | $0.796 \pm 0.092$ | **$0.826 \pm 0.091$** | $0.560 \pm 0.063$ |
| Liver | $0.959 \pm 0.014$ | **$0.964 \pm 0.015$** | $0.882 \pm 0.012$ |
| Spleen | $0.810 \pm 0.103$ | $0.840 \pm 0.077$ | **$0.881 \pm 0.003$** |
| Kidney | $0.867 \pm 0.124$ | **$0.933 \pm 0.062$** | $0.927 \pm 0.009$ |
| Bowel | $0.824 \pm 0.066$ | **$0.856 \pm 0.030$** | $0.705 \pm 0.025$ |
| Bladder | $0.919 \pm 0.077$ | $0.934 \pm 0.029$ | **$0.946 \pm 0.002$** |
| Average | 0.877 | 0.902 | 0.842 |

After the single-organ networks were tested, we tested a network that performed segmentation of all organs simultaneously. Table 2 compares the Dice coefficients between single-organ and multi-organ networks (all pre-trained). The multi-organ network produced significantly higher Dice scores, and lower Dice standard deviations, compared to its single-organ equivalents. The improvement in Dice coefficients was greater than 0.005 with all organs except the parotid gland and liver, which already had very high dice coefficients of 0.948 and 0.959, respectively. Among all organs, the worst measured Dice coefficient was 0.826 for the sublingual gland.

For reference, Dice coefficients measuring the agreement between three human raters (i.e. segmentations from two different physicians and a nuclear medicine technologist) are also included in Table 2. With the exception of spleen and bladder, the Dice values for inter-rater agreement were worse or equivalent than those produced by PSMA-Hornet. One of the largest differences was found with the sublingual gland, where the inter-rater Dice was 0.560, and 0.826 for PSMA-Hornet segmentations.

The recall, precision, and Hausdorff distance metrics for the single-organ and multi-organ networks are given in Table 3. The recall was greater with the multi-organ network for all organs. On the other hand, single-organ networks generally had better precision. In other words, the multi-organ network was more sensitive, but the single-organ networks had on average fewer false positives. The Hausdorff distances were generally smaller for the multi-organ network.

**TABLE 3:** Segmentation quality metrics for single-organ and multi-organ networks.

| Organs | Recall |  | Precision |  | Hausdorff Distance |  |
| --- | --- | --- | --- | --- | --- | --- |
|  | Single-organ | Multi-organ | Single-organ | Multi-organ | Single-organ | Multi-organ |
| Lacrimal | $0.88 \pm 0.06$ | **$0.91 \pm 0.05$** | **$0.93 \pm 0.06$** | **$0.93 \pm 0.04$** | $1.02 \pm 0.07$ | **$1.00 \pm 0.00$** |
| Parotid | $0.95 \pm 0.03$ | **$0.97 \pm 0.02$** | **$0.95 \pm 0.02$** | $0.93 \pm 0.02$ | $2.27 \pm 1.75$ | **$1.92 \pm 1.66$** |
| Submandibular | $0.87 \pm 0.03$ | **$0.93 \pm 0.02$** | **$0.96 \pm 0.05$** | $0.94 \pm 0.05$ | **$1.54 \pm 1.06$** | $1.68 \pm 1.16$ |
| Tubarial | $0.81 \pm 0.09$ | **$0.88 \pm 0.06$** | **$0.88 \pm 0.08$** | $0.87 \pm 0.07$ | $1.79 \pm 0.79$ | **$1.30 \pm 0.50$** |
| Sublingual | $0.79 \pm 0.11$ | **$0.87 \pm 0.11$** | **$0.83 \pm 0.15$** | $0.80 \pm 0.11$ | $1.51 \pm 0.71$ | **$1.43 \pm 0.66$** |
| Liver | $0.96 \pm 0.01$ | **$0.97 \pm 0.01$** | **$0.96 \pm 0.02$** | **$0.96 \pm 0.03$** | $4.74 \pm 3.21$ | **$3.62 \pm 2.80$** |
| Spleen | $0.74 \pm 0.17$ | **$0.80 \pm 0.15$** | **$0.93 \pm 0.06$** | $0.91 \pm 0.05$ | $4.41 \pm 2.71$ | **$4.16 \pm 2.61$** |
| Kidney | $0.80 \pm 0.16$ | **$0.92 \pm 0.11$** | **$0.99 \pm 0.01$** | $0.95 \pm 0.03$ | $3.02 \pm 2.52$ | **$2.17 \pm 2.30$** |
| Bowel | $0.76 \pm 0.11$ | **$0.85 \pm 0.05$** | **$0.91 \pm 0.04$** | $0.88 \pm 0.06$ | $9.12 \pm 5.05$ | **$6.51 \pm 4.26$** |
| Bladder | $0.92 \pm 0.05$ | **$0.93 \pm 0.05$** | $0.93 \pm 0.09$ | **$0.94 \pm 0.05$** | $2.24 \pm 2.03$ | **$1.71 \pm 1.10$** |
| Average | 0.85 | **0.90** | **0.93** | 0.91 | 3.17 | **2.55** |

Examples of organ segmentations produced by PSMA-Hornet in 3 different subjects are shown in Fig. 4, with reference to the original PET images and manual delineations. In general, we found outstanding agreement between the manual and automated segmentations. In large organs with high tracer uptake (liver, kidneys, parotid glands) the manual and automatic segmentations were visually indistinguishable. The largest differences between the segmentations can be seen in the spleen and bowel.

**FIGURE 4:**
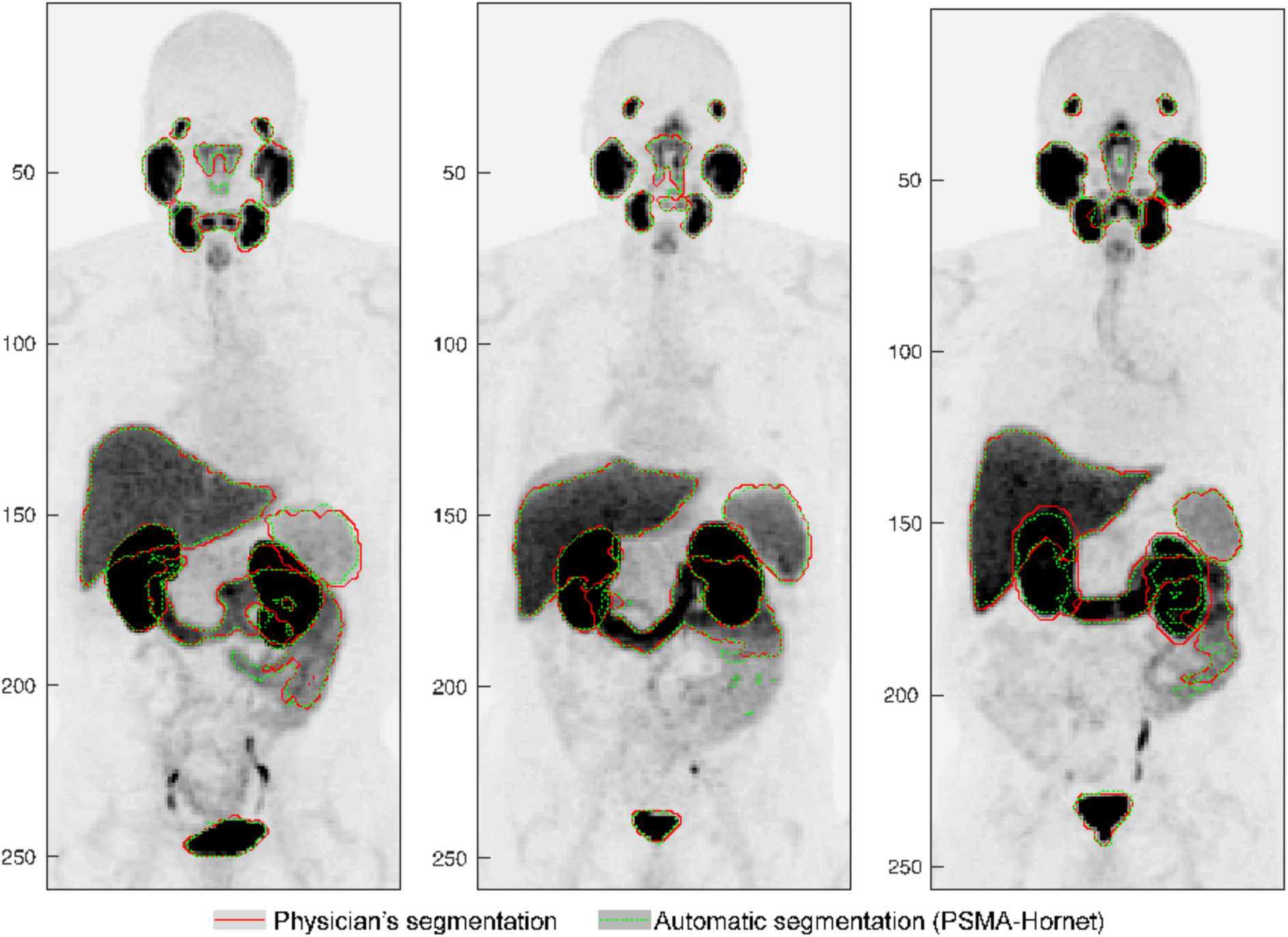
Examples of automated organ segmentations produced by PSMA-Hornet in three different subjects, in comparison to manual segmentations. Maximum intensity projections are shown. Contours of different organs have the same color for visualization clarity.

In some cases, we found that automatic segmentations were more consistent than manual segmentation. For example, in Fig. 4, the automatic contours for spleen follow the organ boundaries more closely than the manual contours. The same can be observed for the bladder, and for kidneys (right panel in Fig. 4). An interesting finding was that urine in ureters was not segmented by the network, despite having very high SUV values.

One of the most common discrepancies between manual and automatic segmentations was the inclusion of high-activity regions in the bowel as illustrated in Fig. 5A (indicated by arrows). However, the observed cases were not necessarily erroneous but rather ambiguous — originating from the inconsistencies in manual segmentations. This can be explained by the convoluted bowel structures with highly variable intensity of uptake and the challenging work required to manually segment them in the absence of reliable automated way to do so. Another less frequent error was mislabeling of high-intensity regions by the network. For example, in Fig. 5A on the right, a group of voxels near the bladder was misclassified as a kidney (indicated by arrow).

**FIGURE 5:**
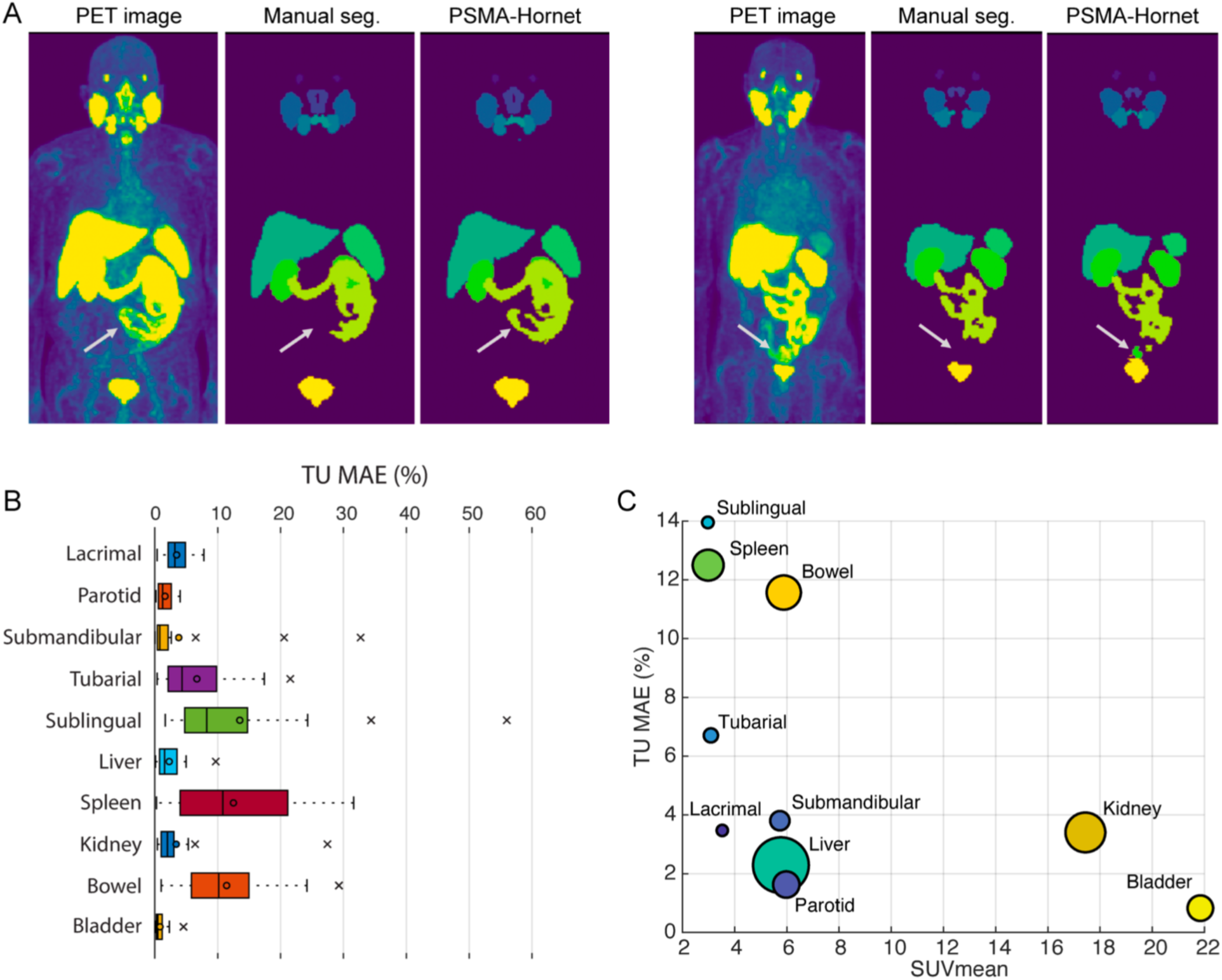
A. Examples of mismatch between manual and automated organ segmentations by PSMA-Hornet. Maximum intensity projections are shown. Arrows indicate regions with significant mismatch. Left: additional bowel voxels were included in the PSMA-Hornet segmentation. Right: a high-uptake region near the bladder was mislabeled as “kidney”. B. The distributions of TU MSEs for different organs. The box plots include data from 30 test samples (combined from 3 test folds). C. The mean TU MSE plotted against the average SUV_mean_ for that organ. The high-risk region combines high SUV_mean_ with high errors of TU estimates (resulting from inaccurate automated segmentation). Size of the marker represents the relative size of the organ.

### Image quantification with PSMA-Hornet

We have determined that PSMA-Hornet produced high-quality organ segmentations, with Dice coefficients ranging from 0.826 to 0.964 for different organs. However, in clinical research and practice, quantitative metrics (e.g. organ volume, SUV_mean_, and TU) are the ones that will be used for diagnosis, outcome prediction, therapy response, etc. We, therefore, evaluated the discrepancy in these metrics introduced by automatic segmentation, as compared to manual delineations.

The relative differences in organ volumes, SUV_mean_, and TU measured with PSMA-Hornet according to Eq. 6 are given in Table 4. The most inconsistent organs in this regard were the sublingual glands, spleen and bowel. The high variability for bowel was likely due to the aforementioned more extensive segmentation tendency of PSMA-Hornet in that organ, while the sublingual glands suffered from small size and low activity values. Overall, the average volume difference (across all organs) was 10.5% ± 6.5%, the average SUV_mean_ difference was 5.3% ± 2.9%, and the average TU difference was 6.0% ± 4.9%.

**TABLE 4:** PSMA-Hornet image quantification errors with respect to manual delineations.

| Organs | Volume MAE (%) | SUVmean MAE (%) | TU MAE (%) |
| --- | --- | --- | --- |
| Lacrimal | $9.51 \pm 4.46$ | $6.40 \pm 3.22$ | $3.47 \pm 1.97$ |
| Parotid | $4.59 \pm 1.62$ | $2.83 \pm 1.02$ | $1.63 \pm 1.10$ |
| Submandibular | $5.15 \pm 5.71$ | $2.96 \pm 2.79$ | $3.80 \pm 8.18$ |
| Tubarial | $10.25 \pm 7.83$ | $3.77 \pm 2.87$ | $6.70 \pm 5.98$ |
| Sublingual | $21.92 \pm 16.77$ | $8.57 \pm 6.51$ | $13.95 \pm 11.60$ |
| Liver | $2.79 \pm 2.17$ | $0.63 \pm 0.41$ | $2.29 \pm 2.26$ |
| Spleen | $20.31 \pm 11.95$ | $9.88 \pm 5.35$ | $12.50 \pm 9.71$ |
| Kidney | $7.76 \pm 10.17$ | $5.70 \pm 9.40$ | $3.40 \pm 4.81$ |
| Bowel | $14.77 \pm 7.36$ | $4.26 \pm 2.12$ | $11.56 \pm 7.04$ |
| Bladder | $8.52 \pm 5.96$ | $8.03 \pm 5.72$ | $0.82 \pm 1.01$ |
| Average | 10.56 | 5.31 | 6.02 |

The difference in mean TU was under 7% for seven out of ten organs, with respect to manual delineations. The detailed distributions of differences in TU quantification for each organ are plotted in Fig. 5B. We observe that for many organs, the TU errors were consistently below 10%. With the exception of one case (sublingual gland), all TU differences were under 35%.

The tolerance level for differences in TU estimation depends on the SUV_mean_ in that organ. It can be argued that for organs with relatively low uptake, the differences in TU estimation are less consequential. In Fig. 5C, the plots of TU MAEs versus SUV_mean_ for different organs demonstrate that organs with the highest differences in TU estimates — sublingual gland, spleen, and bowel — all have relatively low SUV_mean_. On the contrary, organs with high SUV_mean_ — kidneys and bladder — have low TU MAEs. It is worth pointing out that the differences in TU were calculated with respect to manual segmentation, which themselves could be imperfect.

## DISCUSSION

We have developed PSMA-Hornet, a deep neural network to automatically segment 14 organs with high tracer uptake in [^18^F]DCFPyL PSMA PET/CT images. The lowest and highest mean Dice coefficients, which measured the agreement between the automatic and manual segmentations, were 0.826 for sublingual gland and 0.964 for the liver. Automatic segmentations produced by PSMA-Hornet had similar or better Dice scores than those for interrater agreement. In addition, the automatic segmentation of organs like spleen and liver appeared to be more consistent than manual delineations by a nuclear medicine physician. The worst expected error in TU quantification with respect to manual delineation was 13.95% (for the sublingual gland), which is arguably within clinical tolerance. These results demonstrate that high-quality organ segmentations can be automatically obtained for PSMA image analysis tasks, including dosimetry. Our model has the potential to enhance the treatment of metastatic prostate cancer by making image information more rapidly accessible, particularly in light of recent FDA approval of [^68^Ga]PSMA-11 and [^18^F]DCFPyL (*21*) and other rapid developments in PSMA imaging.

In terms of immediate clinical applicability, it would be of interest to develop a similar model for theranostic PSMA-SPECT/CT images. We believe that PSMA-Hornet can be efficiently fine-tuned and applied to lower-resolution PSMA SPECT/CT images, which share the same biodistribution and relative organ uptake. This would allow to calculate precisely how much radiation is being delivered to healthy organs during PRLT. Another possible extension of PSMA-Hornet would be to automatically segment high tumor loads by masking out healthy organs, making our model the first step towards a full-patient segmentation.

We found that using self-supervised pre-training meaningfully improved training convergence, train-to-test generalization, and resulting segmentation quality. The proposed pre-training method does not require any image segmentations or labeling. This can be particularly beneficial in situations where, in addition to a small set of labeled images, there is a large number of images that have not been manually or otherwise processed. We presume that pre-training helped to encode anatomical and functional priors within the network, such as organ shapes, as well as spatial relationships between organs.

Interestingly, a multi-organ (multi-target) network performed better than any single-organ network of a similar architecture. For example, the mean Dice coefficient for kidney segmentation was 0.867 ± 0.124 with single-organ network, and 0.933 ± 0.062 with multi-organ network (Table 2). This is despite the fact that the number of trainable parameter per-target was smaller in the multi-organ network. One possible mechanism for improved performance is the explicit co-activation of output neurons based on the localization and relative size relationships between organs. For example, if spleen is present in a particular target slice, then liver is also likely to be present in that slice. Having multiple prediction targets is also likely to have a regularizing effect on the latent image representation, making it more dense and thus allowing for more robust outputs.

The lowest Dice coefficients with PSMA-Hornet were measured for the tubarial gland, sublingual gland, spleen, and bowel. Interestingly, these organs also had the lowest inter-rater agreement, indicating that human readers and the network were likely challenged by the same image aspects. Difficulties in segmenting the tubarial and sublingual glands were likely related to their small size and relatively low PSMA tracer uptake, making for ill-defined boundaries. The relatively low Dice coefficient for bowel was likely due to the high anatomical variability of that organ, as well as varied uptake patterns (i.e. tracer accumulates selectively in different parts of bowel).

The segmentation quality of PSMA-Hornet could likely be further improved by adding more subjects to the training set or applying data augmentation to the images, such as adding random rotations and scaling. Additionally, heuristic post-processing could be applied to segmentations, e.g. removal of small connected components, or by applying a set of rules that specify the relative positions of organs. We did not apply such post-processing here since our goal was to evaluate the performance of CNNs on their own. Another possible improvement to the network could be the inclusion of multi-organ probabilistic shape priors (*22*).

An important consideration is that human-generated organ delineations, typically used as manual for training, usually include some level of inconsistency. We observed that in some cases, automatic segmentations were more accurate than physician-generated segmentations, particularly in the bowel and spleen. In this sense, the network could handle some inconsistencies (“edge cases”) in the training set, and on the test set, it produced more consistent segmentations compared to a physician. Phantom studies, or studies with consensus segmentations, could help obtain a more objective measurement of segmentation performance albeit without the added value of being representative of the clinically achievable manual work. In future work, it is of interest to investigate how the network handles cases with high tumor load, physiological changes like a bladder with or without diuretic stimulation, and if automated organ segmentation by PSMA-Hornet can also contribute to improved tumor detection and segmentation.

## CONCLUSIONS

We developed and tested PSMA-Hornet, a neural net for fully-automated segmentation of 14 organs with high tracer uptake on PSMA PET/CT images. PSMA-Hornet achieves high-quality segmentations, with the lowest and highest Dice scores of 0.826 and 0.964 for the sublingual gland and liver, respectively. On average, the difference in total uptake quantification in organs was 6.0% ± 4.9% compared to manual segmentations. Self-supervised pretraining and multi-target training were found to improve segmentation quality. The model can provide automatic organ segmentations scalable to clinical PSMA imaging trials and routine radiation dosimetry, where manual segmentation is too labor-intensive. In retrospective studies the model can be used to mine the imaging data and cross-reference it to available patient clinical data, opening up new research and clinical applications.

## Data Availability

All data produced in the present study are available upon reasonable request to the authors

## DISCLOSURES

All authors have no conflicts of interest. This work was supported by the CIHR Project grant PJT-162216, NIH / CIHR QIN grant 137993, and Mitacs Accelerate grant IT18063. Azure Cloud compute credits were provided by Microsoft for Health.

## ACKNOWLEDGMENT

We would like to thank Yixi Xu and Anthony Ortiz from Microsoft’s AI for Good lab for helpful discussions.

## Notes

### Competing Interest Statement

The authors have declared no competing interest.

### Author Declarations

University of British Columbia - BC Cancer Ethics Board (UBC BC Cancer REB) gave ethical approval for this work

